# Airborne magnetic nanoparticles: environmental risk factors for the transmission of SARS-CoV-2

**DOI:** 10.1101/2020.12.10.20247130

**Authors:** C. Martinez-Boubeta, K. Simeonidis

**Affiliations:** Ecoresources P.C., Giannitson-Santaroza Str. 15-17, 54627 Thessaloniki, Greece; Dept. Physics, Aristotle Univ. Thessaloniki, 54124 Thessaloniki, Greece

**Keywords:** particulate matter, iron-bearing nanoparticles, airborne, COVID-19

## Abstract

**Objectives:** To examine the impact of concentrations of ambient fine particulate matter (PM2.5) air pollution on the incidence of COVID-19.

**Methods:** Publicly available data of COVID-19 deaths in March/October 2020 were compared with concentrations of PM2.5 measured in previous years at urban and suburban areas in Thessaloniki. Similar publicly available data of PM2.5 concentrations from Tehran were gathered for comparison. Cross-correlation and Granger causality analysis were performed in order to assess linkage.

**Results:** On the one hand, the mean PM2.5 concentrations in Thessaloniki were significantly higher in the winter, however the magnetic fraction of particulate matter in the autumn is twice its annual average, suggesting that traffic-related emissions alone may not explain the entire variability of PM2.5. On the other hand, it is implied that changes in coronavirus-related deaths follow changes in airborne magnetite, with the correlation between the two data sets being maximized at the lag time of one-month. Further insight is provided by the monthly pattern of PM2.5 mass concentrations in Tehran. We find that air pollution Granger causes COVID-19 deaths (p<0.05).

**Conclusions:** A significant association has been found between PM2.5 values and the impact of the COVID-19 pandemic on a bunch of regions. Reported links between pollution levels, climate conditions and other factors affecting vulnerability to COVID-19 may instead reflect inhalation exposure to magnetic nanoparticles. A hypothesis has been set that ubiquitous airborne magnetite pollution, together with certain climatic conditions, may promote a longer permanence of the viral particles in the air, thus favoring transmission.

**Key messages:** *What is already known about this subject?:*

▸▸ Due to their small dimensions, airborne particles are able to penetrate through inhalation into many human organs, from the lungs to the cardiovascular system and the brain, which can threaten our health. Research has shown that air pollution is an important cofactor increasing the risk of mortality from coronaviruses.

*What are the new findings?:*

▸▸ Evidence exists that the magnetic fraction of PM has modulated the transmission of SARS-CoV-2 in Thessaloniki, and potentially in any other region in the world.

*How might this impact on policy or clinical practice in the foreseeable future?:*

▸▸ Policymakers should take care not to overestimate the effect of social distancing interventions and should consider the impact of air pollution in current or future epidemic waves.

## INTRODUCTION

A large body of literature has shown that long-term exposure to high levels of air pollution can cause premature death among the elderly and more than 3 million cardiopulmonary mortalities per annun [1]. In this context, it has been also suggested that air pollution is an important cofactor increasing the risk of mortality from coronavirus. For instance, analysis of the first severe acute respiratory syndrome coronavirus SARS-CoV-1 outcomes in 2003 demonstrated that in heavily polluted regions the risk of dying from the disease was >80% higher compared with areas with relatively clean air [2]. Furthermore, a correlation -but not necessarily a causal link-between NO_2_ levels (a pollutant produced mostly by diesel vehicles) and COVID-19 cases has been recently reported [3], while another study supports the hypothesis that the viral infectivity is higher in hinterland cities rather than coastal areas due to airflow conditions that prevent dispersal of air pollution [4].

Nonetheless, while there has been ample evidence for a relationship between long-term air pollution exposure and the severity of COVID-19 outcomes [5,6], one aspect that has yet to be described is the effect of seasonal variances on excess mortality from COVID-19.

Apart from mass concentration and particle size distribution of aerosols, oxidative potential has been suggested as possible driver of both the chronic and acute consequences on human beings [7]. In this regard, high oxidative properties of PM have been attributed to the high iron levels in vehicles emissions [8,9]. This adds to the intrinsic redox activity of particles originating from wood burning [10], which probably arises from the presence of magnetite and other co-associated metal-bearing nanoparticles in soot and smoke [11,12,13].

At the time of writing, Greece is bracing for what seems to be a second wave of the COVID-19 pandemic after a surge in cases during late summer, especially in northern Greece where most victims are count in hospitals of Central Macedonia. It somehow replicates the seasonal variations of anthropogenic magnetite reported in a forthcoming paper [14]. Consequently, we aimed to verify whether PM2.5 may exacerbate the impact of pandemic waves.

## METHODS

Estimation of exposure of inhabitants of Thessaloniki to airborne magnetite was determined by superconducting quantum interference device (SQUID) magnetometry as described elsewhere [14]. Briefly, cumulative 24-hour samples were collected on PTFE PM2.5 (referring to particles smaller than 2.5 μm) filters, using a vacuum pump, operating at a flow rate of ∼2 m^3^/h. Seasonal arithmetic mean concentrations were calculated over the period from February 2015 to October 2018. The routine analysis comprises gravimetric monitoring with a precision microbalance (±1 μg); trace element quantification using scanning electron microscopy (SEM) and energy dispersive X-ray analysis (EDS); crystal and morphological characterization by transmission electron microscopy (TEM). The magnetic fraction was obtained after rinsing the filters in a mixed solution of acetone and polyvinyl alcohol under the influence of an externally imposed magnetic field gradient by means of a NdFeB magnet. Complimentarily, magnetic nanoparticles were also detected in swab specimens collected in late autumn 2019 from the nasal mucosa of patients in the Otorhinolaryngology Clinic of the General Hospital “Papageorgiou”. All patients gave written informed consent before study inclusion.

To examine the potential association between PM2.5 levels and spread of COVID-19 in big cities located in different geographical areas, statistical analysis was performed on data from Tehran, based on Nabavi *et al*. [15]. The monthly pattern of PM2.5 mass concentrations during the period (2011–2016) was compared to confirmed deaths from COVID-19 in Iran, as reported by the “Worldometer” website, updated with daily frequency [16]. The time-series data were interpolated to evenly spaced observations [17]. Lead-lag relationships were investigated by cross-correlation analysis of monthly data during the period from March to October. Granger causality was computed by testing the null hypotheses in the Free Statistics Software from Wessa.net [18].

## RESULTS

We build on the observation of magnetic traces of in vivo human nasal swab samples [14]. The magnetically responsive dust is made of aggregates in which particles with rounded morphologies and a mean size around 15 nm appear basically made of magnetite (and minor contribution of Fe^3+^ rich shell and substituted heavy metals such as Cr, Mn, Co or V).

Our findings suggest that, given their tiny size < 5 μm, these combustion-derived particles remain airborne for a long time [19], and can penetrate the airways all the way down to the alveolar space [20]. Furthermore, inhalation of PM may trigger susceptibility to respiratory infections, such as the SARS-CoV-2, since nasal surfaces are suggested as initial loci of disease [21]. According to this hypothesis, magnetite could then tentatively offer a Trojan horse docking for virus infectivity via the Fenton-mediated hydroxyl radical production. Indeed, recent studies revealed that iron oxide nanoparticles interact efficiently with the SARS-CoV-2 spike binding proteins [22].

In this framework, we analyzed whether seasonal variations of airborne magnetite has positive qualitative effects on coronavirus mortality. Results for Greece are shown in Fig. 1. Two main conclusions can be drawn from this chart: first, given the current epidemiological situation and with regards to the re-opening of schools on 14^th^ September, the decision was not insignificant at all. The figure above shows that the country was making progress on bringing down the curve of new coronavirus deaths until when it was necessary for families to return to big cities from summer vacations, and consequently air pollution peaked. It adds to evidence that adolescents can seed clusters of COVID-19 cases [23].

**Fig. 1.**
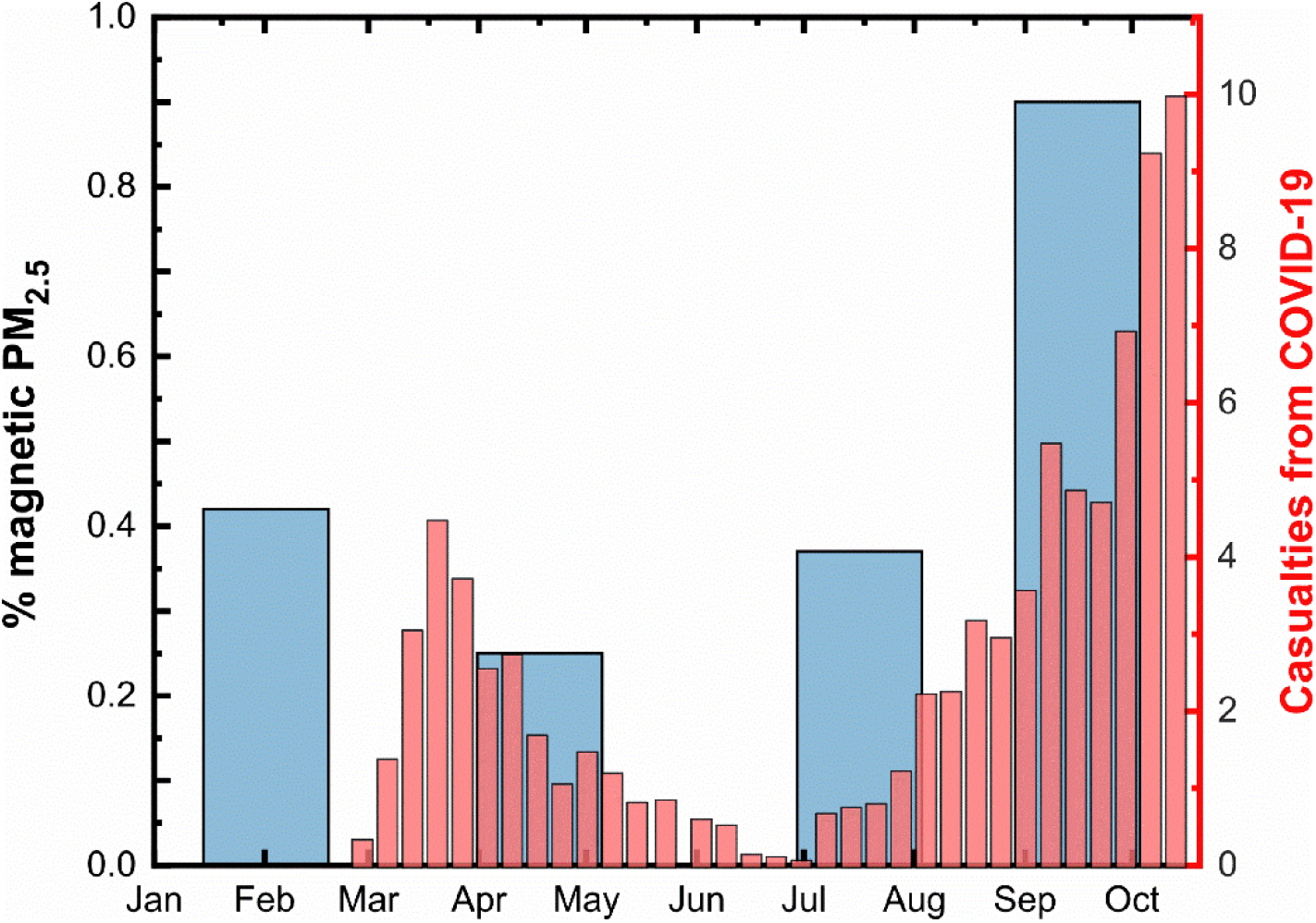
The disease progression timeline and the seasonal variability of the airborne magnetite pollutant. The thin red bars represent the 7-period moving average for new daily confirmed deaths from COVID-19 in Greece (data retrieved from Worldometer database). Note that there is about a four weeks delay from first symptoms after infection to death. The thick blue bars depict the percentage of magnetic material estimated from PM2.5 observations at urban site in Thessaloniki, representatively for the years 2015-2018.

Secondly, the air pollution conditions may had been favorable for the virus circulation since late September 2019, much earlier than the first reported cases. We note that WHO declared the pandemic on 11^th^ March 2020. Officially, a 38-year-old woman who returned to Thessaloniki from Milan by air on 23^rd^ February 2020 became Greece’s first coronavirus. Though, a growing body of evidence suggest that the new SARS-CoV-2 virus had already been circulating unnoticed in the community a few months before the first reported case in Wuhan City, China, by December 2019 [24].

It is also worth noting that during the winter and spring times the mean PM2.5 concentration in Thessaloniki exceeds the European ambient air quality standard of 25 μg/m^3^ [14]. It would mean that the attributable fraction of COVID-19 mortality due to the long-term exposure to ambient fine particulate air pollution could be as high as ∼50% [5]. But when this analysis is applied to the magnetic percentage attributed to air pollution from anthropogenic sources that peaks during the autumn, it potentially explains the rise in incidence of the second wave of the pandemic outbreak. Magnetic particulate matter shows maximum values during autumn months (0.8 % wt.), compared to <0.4% in spring, meaning that doubling the concentration of airborne pollutants leads to a ten-to 20-fold increase in the number of deaths linked to COVID-19. This goes in line with other studies, which have found that the association between the pollutant and the health outcome is log-linear [25,26]. Consequently, our research strongly suggests that magnetic parameters can be used as an efficient proxy to assess the urban atmospheric quality and the impact of COVID-19 in Thessaloniki, and potentially in any other region in the world. Therefore, further investigations focusing on the development of COVID-19 outbreaks over highly polluted areas are encouraged.

For the while let us, for historical reasons, focus on Iran since the ancient Persian and Greek cultures intermingled when Alexander the Great conquered most of that region by 330 BC. In addition, interest arises from the fact that, when it comes to number of new deaths per day, the third wave of coronavirus in Iran is the worst yet (Fig. 2).

**Figure 2.**
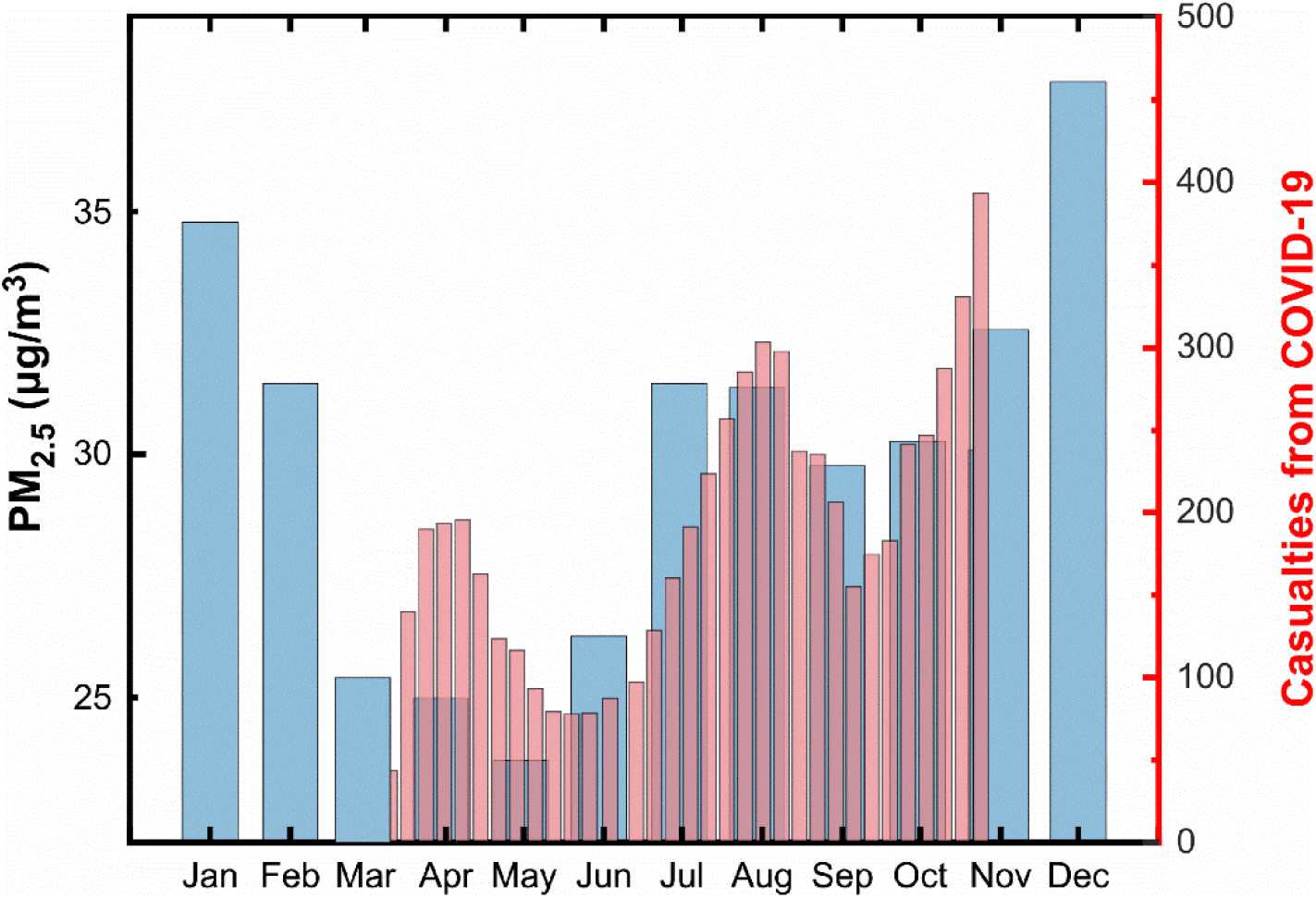
Two or more ‘camel humps’ in Iran. The thick blue bars depict the monthly pattern of PM2.5 mass concentrations during the period (2011–2016) in Tehran. The thin red bars represent the 7-period moving average for new daily confirmed deaths from COVID-19 in Iran, as of 28th October 2020.

On this matter, Iran as a developing country is facing severe problems in terms of air quality. Urban air pollution due to PM2.5 in Tehran and other regions of the country has been reported by many researchers [15,27,28,29]. Accordingly, most cities in Iran have PM2.5 concentrations above the WHO air quality guideline value, which in turn are magnified by frequent desert dust storms during the summer months as temperatures rise and rainfall reaches a minimum [30]. However, the highest amounts of PM2.5 are normally recorded during winter. The increase of PM concentration in the cold period of the year is due not only to the increased emissions from additional sources, such as domestic heating, but rather also to the decrease in the thickness of the mixing layer. These episodes have had notable health effects. As such, the highest number of deaths due to exposure to fine particles was estimated to occur in Iran’s largest city, Tehran, where motor vehicles play a major role. Other episodes that may be linked to human activity include the significant drop of PM in the spring caused by Tehran’s minimal traffic flow during Nowruz holidays.

The figure above depicts that the coronavirus casualties follow the seasonal variation of air pollution closely, except that the PM2.5 waves start sometime early. This plot is of special interest since the data can be used to validate the causal association between pollution and COVID-19 severity. Techniques exist for establishing the direction of causality and have previously been used primarily in fields such as econometrics [31]. For instance, to say “one variable X Granger causes another variable Y” means that, by using past values of both X and Y, we can better predict future values of Y than by using only past values of Y. So the prediction of Y is significantly improved by including X as a predictor. Let Y = *daily deaths* be the potential outcome in the population of a country exposed to X = *pollution concentration*. We computed Granger causality by testing the null hypotheses of X does not Granger-cause Y and vice versa. The results of these tests are reported in Table 1.

**Table 1.** Iranian values of the constructed proxies for PM2.5 pollution (X) and the health impact (Y) on COVID-19, and both cross-correlation and Granger causality summaries identifying significant causal relationships between X→Y. Significance level **p < 0.05.

| X | Y | lag | $\rho(Y[t], X[t+\text{lag}])$ | Granger-Causal | $Y=f(X)$ | $X=f(Y)$ |
| --- | --- | --- | --- | --- | --- | --- |
| 25 | 65 | -4 | -0.12 | F | 8.33 | 0.04 |
| 23.7 | 140 | -3 | 0.10 | p | 0.04 | 0.86 |
| 26.3 | 74 | -2 | 0.53 |  |  |  |
| 31.5 | 98 | -1 | 0.71** |  |  |  |
| 31.4 | 199 | 0 | 0.51 |  |  |  |
| 29.8 | 223 | 1 | 0.16 |  |  |  |
| 30.3 | 159 | 2 | 0.04 |  |  |  |
| 30.1 | 262 | 3 | -0.27 |  |  |  |
|  |  | 4 | -0.32 |  |  |  |

The correlation between the two data sets is maximized at the lag time of one-month, which is reasonable given that there are significant delays between infection, the onset of symptomatic disease, and recovery or death; people who died this week were most likely infected as far back as a month ago [32]. This implies that the changes in coronavirus-related deaths follow the PM2.5 changes. Furthermore, according to the p-values in this table, PM2.5 values provide statistically significant information about the future values of deaths: the p-value of 0.04 means the null hypothesis that X does not cause Y can be rejected. On the other hand, the p-value of 0.86 allows us to accept the null for X = f(Y). The fact that first hypothesis was rejected and second was not means that X can be used to forecast Y. Moreover, we have conducted a sensitivity analysis by using mid-month values on the 15^th^ day, and log-transformed data, and the results remained robust and consistent. Consequently, it is safe to conclude that a portion of the variability of the clinical syndrome of COVID-19 might be affected by environmentally driven variance of nasal infectivity.

## DISCUSSION

The relationship to COVID-19 mortality in Iran is not unexpected, considering the significant association between patterns of PM2.5 mass concentrations and cases of cardiovascular mortality. For example, a study of the 2008-2017 period showed that 2009 and 2010 were the most polluted and unhealthy years, the lowest being 2014, coincidentally during the minimum and maximum of solar cycle 24, respectively [29]. Incidentally, note that a small amount of iron oxides arrives as cosmic flux [33]. This relationship has significant practical implications deserving of further research.

Despite their diversified sources of data and the methods of reconstructions, the strikingly high congruence among the PM2.5 pollution and the COVID-19 impact records warrants the reliability of the analysis. Further support for this causal interpretation also comes from literature on associations of daily pollution variations of PM2.5 and black carbon –while the same is not true for nitrogen dioxide NO_2_– on daily deaths in the Boston area, U.S. [34]. It is also noteworthy that similar Granger causal relationships have been reported between daily values of PM2.5 pollution and new daily COVID-19 infections in Emilia-Romagna, Italy [35]. The explanation is particularly interesting as the phenomena seem to be global in scale. This may have possible implications for the evolution of pandemic coronavirus in distant places of the Greek diaspora as far as California and Australia (see Supporting Information). Therefore, the association between PM2.5 and deaths during this pandemic is almost certainly causal. In this regard, we note that there is growing evidence that indoor airborne transmission due to droplets <2µm is the primary driver of the global COVID-19 pandemic [36]. Hence, the study of PM2.5 may be allied to some truths in medicine when battling COVID-19 pandemic.

The present study has several limitations that should be considered when interpreting its findings. First, our causal modeling is designed to handle pairs of variables, in which only linear relationships between predictor (pollution) and target variable (casualties) are considered, thus it may suffer from a typical limitation when a third variable is engaged. For instance, variation of meteorological factors (such as temperature, relative humidity, rainfall and prevailing winds) confines the vertical distribution of air contaminants, but also modulates human activities such as commuting choices and leisure occupation, and would subsequently impact the relationship between PM and the spread of infections. Consequently, the second issue is the potential of missing other important confounding factors (Supporting Information). A third limitation of this study is missing data. The estimate results obtained here are preliminary and must be treated with caution when analyzing, as in our case, short time series. This adds to the fact that the environmental study was performed at the city level, and inference of the epidemic dynamics at the nationwide level is dubious. But overall, limitations in data availability on the occurrence of magnetite in the atmosphere remain the main obstacle to conducting conclusive studies on this topic. Given the above considerations, future studies are needed involving a larger number of observations.

Final mention must go to the fact that our results are provisional, based on epidemiological data collected up to the end of November 2020, and a comprehensive evaluation will need to follow after the COVID-19 pandemic. In this regard, it is to be seen whether our initial conclusions remain appropriate and would be relevant to the public health response to future outbreaks.

## Data Availability

All data used in this study are available from public repositories, in particular the Worldometers Database (https://www.worldometers.info/coronavirus), referenced studies and/or from the corresponding author on request.

## Contributors

Both authors conceived of the idea for the study. Both authors designed the study. CMB conducted data analysis and drafted the manuscript, which KS critically reviewed.

## Patient consent for publication

Not required.

## Provenance and peer review

Not commissioned; externally peer reviewed.

