## Supplementary Information for "Airborne magnetic nanoparticles: environmental risk factors for the transmission of SARS-CoV-2"

### 1. ON THE RELATIONSHIP BETWEEN AIR POLLUTION AND COVID-19

More than three decades ago, it was demonstrated in randomized control trials that mice exposed to higher PM concentrations experienced increased mortality when infected with a common strain of the influenza virus [1], as it is also the case for bacterial infections of the lower respiratory tract [2]. From there on, there is mounting evidence of increased susceptibility to respiratory infections from exposure to air pollution [3,4,5].

Furthermore, a hypothesis has been set that air pollutants, together with certain climatic conditions, may promote a longer permanence of the viral particles in the air, thus modulating the spreading and virulence of COVID-19 [6,7,8,9,10]. It might very well happen that coronaviruses use pollution particles as a Trojan horse. Indeed, both the airborne PM and the SARS-CoV-2 virus enter the body via the upper airways and share common pathophysiological mechanisms of oxidative stress and immune responses [11,12]. This parallels the well-established pathogenic role of the microbial component of PM [13]. Onto this we add the increased mutagenicity risk of PM extracts related to the presence of magnetite [14].

The underlying mechanisms of the toxicological impact of particulate matter are not yet fully understood, but a widely accepted hypothesis is that they are capable of generating reactive oxygen species (ROS) and thus inducing oxidative stress at the cellular level. The most representative example of ROS is the hydroxyl radical catalyzed by ions of iron, copper and other metals. In this context, Kocbach *et al.*[15] have reported that wood smoke may have a similar inflammatory potential as traffic-derived particles, both affecting the release of cytokines TNF-α, IL-1β and IL-8 in monocytic cells. Noteworthy, in COVID-19, TNF-α has been a prominent feature of patient deterioration, increasing in ICU patients, as well as cytokines IL-1β and IL-8 [16]. The report by Maher *et al.*[17] provides information on this subject by establishing that the level of exposure to PM2.5, and to magnetite particles, originating from open fires is similar to and indeed might exceed that from roadside sources. The source of this ubiquitous and abundant magnetite contamination in airborne dust is probably phytoferritin [18], which is also consistent with the findings presented here.

Regarding Greece, in a previous study [19] we showed that the mean winter PM2.5 concentrations on weekends were higher than on weekdays in Thessaloniki area, but the reverse is true when it comes to the magnetic fraction of particulate matter. Thus suggesting that traffic-related emissions alone cannot explain the entire variability of PM2.5. This is because the reduction in road transport and industrial activities were counter-balanced by an increase of PM emissions from the activities at home (e.g. biomass burning in domestic heating) during late autumn to early spring. Our results are in line with a recent paper by Dobson & Semple [20] that examined the impact of COVID-19 lockdown restrictions on air pollution and suggests that personal exposure to PM2.5 may result in social health inequalities since lower income people are likely to life in smaller homes leading to higher PM2.5 concentrations indoors. This could tentatively explain the evidence of declining susceptible-infectious transmission probability in larger households [21,22], an unexplained factor that was previously reported for the Spanish Flu [23]. Recent analysis and modeling results provide partial support to the hypothesis by showing that disease transmission occurs in a closed ambience (e.g. the workplace, schools, at home, restaurants, and in hospitals) [24,25] and there are higher infection rates among disadvantaged racial and socioeconomic groups [26].

All this provides the main framework that gives shape to our investigation. And allow us to conclude that air pollution may have favored the spread and virulence of COVID-19 in various regions of the world, which we will explore next.

### 2. LESSONS FROM ABROAD AND THE PAST: DRAWING ON EXAMPLES OF INDIA, AUSTRALIA AND UNITED STATES

#### Fighting the Spanish flu pandemic in the U.S. in 1918

We believe that historical analysis is another powerful source of information to test the feasibility of our hypothesis. It remains a source of surprise how closely the present circumstances resemble those of the earlier ages. Much like today, past pandemics have tended to come in waves, the first mild wave in the spring followed by a larger fall wave that would eventually disappear even without significant human intervention [27].

A paradigmatic example is offered by the abundant information regarding the timing, severity, and excess mortality of the 1918 Spanish flu pandemic [28,29]. A variety of interventions were available in 1918, from wearing surgical masks to ‘‘social distancing’’ measures that ranged from closing schools and prohibiting public gatherings to isolating sick people. Most of the interventions then available are identical to the measures that are in used today. Nonetheless, past public health interventions are estimated to have achieved only a moderate (< 10%) reduction in mortality [30]. Furthermore, there was also some local intrinsic variation in overall mortality, as city-specific per-capita excess deaths in 1918 was significantly correlated with prepandemic mortality. These facts all agree with our hypothesis on the link between magnetite air pollution and viral infection outcomes. In fact it probably has its roots in pollution exacerbated by the use of coal, as cities that used more coal-fired electricity generation experienced tens of thousands of excess deaths in 1918 relative to cities that used more hydroelectricity [31]. We refer the reader to the body of the paper for discussion on the relationship between soot carbon and magnetic particles.

Although air quality has improved dramatically over the past 100 years in the U.S., urban air pollution remains a major problem. In regards to the 1918 pandemic, the relative ambient concentrations of atmospheric black carbon in Chicago, and other cities in the U.S. Rust Belt such as Detroit and Pittsburgh, peaked during the first decades of the 20$^{th}$ century [32]. Nowadays, the mean annual PM2.5 concentrations are still generally higher in eastern U.S. than in the semi-arid western United States, which is associated, for instance, with a much higher PM mortality burden and hazard ratios of Parkinson's disease [33,34]. And, we believe, could also partially explain the divergent temporal dynamics and severity of the COVID-19 outbreaks (e.g. New Jersey vs. California; data retrieved on 28$^{th}$ October 2020 from https://www.worldometers.info/coronavirus/country/us).

Continuing with the discussion on California's response to the COVID-19, we note that big fires use to appear in a spatially and temporally distinct pattern, and the controlling climate factors diverge: summer fires tend to burn between June and September, while wind-driven Santa Ana fires occur mostly in October through December [35]. Furthermore, summer temperatures well above-normal and extreme drought [36], has made 2020 the largest wildfire season recorded in California's modern history. In view of the foregoing observations, we believe it is safe to conclude that the bimodal fire pattern also carries implications for efforts to predict future COVID-19 outbreaks, since the surge in daily new cases and mortality appears to follow the same course.

In the past, like today, the economic costs were substantial, almost 6 percent of total U.S. GDP in 1918, and the estimated mortality attributed to pollution was above 20% [31] Therefore, further investigations focusing on the development of COVID-19 outbreaks over highly polluted areas are encouraged. For the while let us focus on India.

**How far has India succeeded in containing the pandemic?**

Many regions in the world are immersed right now in a very challenging second wave of the pandemic. On the contrary, India's COVID-19 casualties have declined rapidly since mid-September (data retrieved on 28$^{th}$ October from https://www.worldometers.info/coronavirus/country/india). The total death toll stands below 150,000, about half that of the U.S., despite being India a developing nation with roughly one-third the size of the United States and four times its population. Explanations for the low death rates are lacking. And most of the models built on epidemiological theory are of little help in this area [37]. For instance, the COVID-19 outbreak in India was predicted to be controlled around the end of May 2020 [38].

Thus, we tentatively turn our attention to linkages between meteorology and pollution. First, we note that India received "above normal" monsoon rainfall this year, recording the second highest precipitation in the last 30 years, which may have helped to keep the number of COVID-19 casualties down. Monsoon arrived in Kerala on 1$^{st}$ June, and withdrew from west Rajasthan and Punjab on 28$^{th}$ September. Second, India, though physically located in northern hemisphere, has distinct seasonality of respiratory diseases. For instance, results of five years surveillance during 2009–2013 revealed influenza circulation primarily from June-October [39], i.e. during the rainy/monsoon season. In states like Maharashtra, Chennai and Vellore, located at the south, there is usually also a second surge during the winter. Third, despite air pollutants vary strongly between regions and seasons, we can naively assume the highest PM2.5 values (> 100 µg/m$^3$) are observed during winter and lowest during the monsoon period because of the washout effect of particles from the atmosphere [40]. This adds to stubble burning episodes, which occur every year in October and November in the Punjab and Haryana states, and affects the whole northern India. Impact of biomass-burning to PM2.5 decreases gradually onwards and, concurrently the relative effect of anthropogenic emissions from vehicular traffic increases to 95% or more during

December [41]. Overall, this widespread PM2.5 results in a considerable impact on life expectancy, with a nationwide estimate of about 47 premature deaths per 10k population in 2011 [42].

In the meanwhile, the Government ordered a nationwide lockdown from 24$^{th}$ March 2020 that gave path to certain relaxations from 20$^{th}$ April through five phases of containment to today. We therefore foresee that as the gradual unlock commenced, air quality will also start deteriorating by the end of October. It is to be seen whether a second wave of COVID-19 infections will follow the rationale portrayed above, but chances are that the number of daily new deaths will peak again during the turning of the year.

**Implications for Australia**

Following with the observed planetary-scale repercussions of the linkage between aerosol events and related spreading of SARS-CoV-2 virus, we note that for northern Australia, the peak fire period is throughout winter and spring, while for southern Australia, the bushfire season peaks in summer and autumn. Although specific weather situations influence the size and intensity of fires. For instance, 2019 was the warmest December on record Australia-wide, and large areas of Australia had their highest accumulated Forest Fire Danger Index [43]. And so the 2019–20 bushfires were unprecedented. It probably helped in the exceptional hemispheric-scale perturbation of the stratosphere following the overshoot of combustion products around the turn of 2020 [44]. And despite the smoke was dispersed across all of the southern hemisphere by the prevailing westerly winds, it returned back like a boomerang and sedimented to lower altitudes during the following months of pandemic. On the other hand, PM2.5 levels are higher in the cooler months of the year, from May to October, due to a combination of woodsmoke from domestic heaters and vehicle/industry emissions [45]. Consequently, building on comparison of data available on COVID-19 cases (https://www.worldometers.info/coronavirus/country/australia), some evidence exists that the fire-derived PM may have modulated the transmission of SARS-CoV-2 in Australia.

In this regard, the concept of wood smoke-specific toxicity to immune pulmonary macrophages has been suggested previously. Franzi *et al.* [46] showed that wildfire particulates were four times more toxic to macrophages on an equal weight basis than other types of PM. As mentioned above, a suggested mechanism to explain these effects is iron oxide-mediated oxidative stress [47].

Fires are predicted to pose more of a threat over the coming months. It is to be seen whether the ongoing La Niña, a seasonal climate driver typically associated with above-average rainfall across most of the northern Australia in the December 2020 to March 2021 period, will help to maintain the deaths number low.

### 3. LIMITATIONS OF THE STUDY

Given these results and prior studies, the association between seasonal patterns of PM2.5 and deaths during this COVID-19 pandemic is almost certainly causal, although the physical mechanism remains unknown. We suggest that attention should be paid to airborne magnetite in the sequence of cause-and-effect of a standard Granger causality approach.

Nevertheless, we acknowledge that the study design has limitations related to the potential of missing other important confounding factors. For example, despite the role of the meteorological parameters is quite evident in this study, they are not quantified. On the one hand, the presence of magnetite in PM extracts has been previously shown to strongly correlate also with meteorological data, in particular, the number of sunny hours and the relative humidity [48]. On the other hand, questions remain as to whether weather factors such as temperature, precipitation, and ultraviolet light can influence the spread of the SARS-CoV-2 virus [49,50,51,52,53]. Indeed, past pandemics seem to have ridden along on conditions such as war, famine, and unfavorable weather [54]. Nonetheless, it has been shown that weather alone could not explain variability in the virus's spread [55].

Neither is clear whether other pollutants such as nitrogen dioxide ($NO_2$) and sulphur dioxide ($SO_2$) may contribute to the risk of such respiratory infections. In addition, direct data linking other heavy metal exposure (e.g. As, Cd, Hg, and Pb) and COVID-19 severity are lacking, although reduction in heavy metal emissions may significantly reduce stress and inflammation both of which are known to increase the risk of chronic diseases.

In this regard, higher historical PM2.5 exposures are positively associated with higher COVID-19 mortality rates [9], but exposure to PM2.5 alone does not fully characterize the toxicity of the atmospheric mix. For example, analysis of air quality in Europe from 2000 to 2017 [56], show that premature deaths attributable to PM2.5 expressed as the number of years-of-life-lost per 100k inhabitants follow Serbia (1920) > Bulgaria (1860) > Greece (1170) > Italy (910) > Germany (720) > Spain (550) > UK = Denmark (490) > Portugal (470) > Ireland (250) > Norway (230), data rounded to the nearest ten, i.e. the largest impacts are observed in central and eastern European countries whereas the smallest relative impacts are found in countries situated in the north-west of Europe. Which is not what is seen clinically in COVID-19. And we also lack an explanation for the fact that past years' seasonal patterns of pollution are able to account for much of the variability of the COVID-19 incidence rates despite the amelioration of air quality following lockdown. Surprisingly, some other recent works [57,58] have concluded that COVID-19 deaths positively affected environmental quality by reducing economic and social activities, meaning that, through various lockdown measures, PM2.5 emissions were also reduced.

Consequently, a refinement of the potential relationship between pollution and pathogen transmission will require additional tracing of magnetite nanoparticles in the atmosphere. Unfortunately, the number of cases in which source identification of airborne particulate matter composition is provided, and particularly the iron content, are scarce. Which makes direct comparison with the results in our study difficult. But luckily there are at least two exceptions to this [59,60], wherein the average seasonal concentrations of Fe roughly coincide with our interpretation of magnetic monitoring of PM2.5.

Similarly, other variables capturing demographic and lifestyle features were not assessed; a non-exhaustive list of risk factors for COVID-19 susceptibility and severity [61], includes: obesity, diabetes, cardiovascular diseases, smoking prevalence, social density, age distribution, etc. For example, there is evidence to suggest an important relationship between cardiorespiratory fitness and health outcomes in patients suffering from COVID-19 [62]. In this regard, cardiovascular risk factors are also in relation to elevated PM2.5 concentrations [63]. Up to now, the COVID-19 mortality rate has been estimated to be ~ 4% in symptomatic cases [64], the most common comorbidities are hypertension (57%), obesity (42%), and diabetes (34%) [65]. Although care is needed to ensure that those are specific predictors of the risk and not simply markers of poor health in general. For instance, the findings of a large international prospective study [66], suggest that wealthier countries, in terms of gross domestic product GDP per capita, had an increased number of COVID-19 deaths per million population, secondary to increased prevalence of obesity and the elderly. Although quite unexpectedly, a higher prevalence of smoking within a population seems to decrease the number of critical cases. Regarding the impression that the intake of nicotine or other constituents in tobacco may affect COVID-19 mortality [67], a statistically significant negative association between smoking prevalence and the incidence of COVID-19 across 38 European nations has been recently reported [68]. At one extreme, about 40% of the adult population of Greece smokes, but it has one of the lowest prevalence of COVID-19 cases per million people in Europe. On the other side of the spectrum are wealthier nations from the western part of Europe (e.g. UK, Sweden) with lower smoking prevalence but higher number of COVID-19 cases and deaths per million people. Nevertheless, little is known about the biological and physicochemical mechanisms that may explain in part this relationship. But it has been demonstrated that cigarette smoke impairs chemokine expression [69,70]. These results highlight the importance of macrophages in generating an appropriate immune response. In this regard, the identification of genetic factors that modulate respiratory virus pathogenesis, such as the CCR5-Δ32 mutation [71,72] that plays a crucial role in controlling memory T-cell recruitment [73], could perhaps shed some light on this. And also aid in understanding the so-called Iberian Peninsula paradox, for close geographical regions with similar socioeconomic conditions do have completely different mortality rates.

While further analysis of this intriguing observation is beyond the scope of our paper, we speculate that two non-exclusive hypotheses might account for key aspects of SARS-CoV-2 prognosis: (1) transmission is mediated by airborne particles directly infecting the lung or (2) the nose is the initial site of infection, followed by translocation of the viral inoculum into the brain. In support of the second hypothesis, we note there is ample evidence that COVID-19 is capable of infecting the central nervous system and cause brain damage even in patients with mild respiratory symptoms [74], as was also the case for the previous deadly coronavirus, SARS-2003 [75]. This adds to the olfactory dysfunction in COVID-19 patients [76], suggesting that nasal infection may contribute to the high penetrance of viruses in various brain regions [77]. Once again, this may be fostered by iron oxide nanoparticles that enter the brain directly by crossing the olfactory unit [78].

In considering the first hypothesis, one potential mechanism is by altering the macrophages clearance, cytokine secretion and ROS, which play a critical role in the pathogenesis of many inflammatory conditions [79], in line with a previous study reporting in vivo magnetite particle migration and lung clearance in smokers and nonsmokers [80].

Altogether, the iron content in air pollution is likely to affect critical biological processes where iron participates, and lead to oxidative stress and inflammation. For instance, it is to be noted that hyperglycaemia has emerged as an important risk factor for death in hospitalized COVID-19 patients [81], in which iron plays a crucial role in protein glycation [82]. Alterations that viciously aggravate the release of pro-inflammatory cytokines that ultimately lead to reduced phagocytic efficiency

[83]. There is also a growing body of evidence correlating oxidative damage to disease states that conduce to overexpression of ferritin (a $Fe^{3+}$ storage protein) [84]. In this regard, Covid-19 patients with elevated levels of ferritin are much more likely to experience more severe symptoms and possibly increased mortality rates [85,86]. This may link to the fact that fidelity of single-stranded positive-sense RNA replication, just like the coronavirus family, correlates with divalent cations concentration [87]. Last, along with the increased oxidative stress, increased ischemia and thrombosis have also been reported in humans after only 1 hour of exposure to traffic particles [88]. Therefore, it is envisaged that PM could account for most of the typical complications of COVID-19.

This complex issue definitively deserves further investigation and we believe that magnetic tracking of airborne particulate matter might be also a method to signal and prevent the spread of coronaviruses supporting the public health system. As a matter of fact, the rapid short-term pain relief after the 2020 spring nationwide lockdowns, adopted to reduce human interaction, was also assisted by the sizeable improvement in air quality across the world [89,90].
